# “Go and bring your husband”: a COM-B guided qualitative study on the barriers to male involvement in antenatal care in Bamenda Health District, Cameroon

**DOI:** 10.1101/2024.02.13.24301733

**Authors:** Lily Haritu Foglabenchi, Tanya Marchant, Heidi Stöckl

## Abstract

**Background:** Increasing access to and utilization of skilled care during pregnancy and child birth can significantly reduce maternal and infant morbidity and mortality. Male involvement can positively influence utilization but resource limited settings like Cameroon encounter obstacles in engaging men in maternal and child health services. The aim of our study was to identify contextually relevant barriers to male involvement in antenatal care attendance to inform the development of an intervention that is aimed at promoting male involvement in maternal and child health in Cameroon.

**Methods:** This study used a qualitative design with qualitative methods that draw on 68 semi-structured interviews and three focus group discussions with pregnant women, male partners and health workers. Both interviews and group discussions were audio-recorded, transcribed. Guided by the Capability, Opportunity and Motivation (COM-B) model of behaviour and Theoretical Domains Framework (TDF), we analyzed data using directed content analysis, followed by inductive thematic analysis.

**Results:** Our findings suggest that male involvement in antenatal attendance in Bamenda Health District is under the influence of six multidimensional factors: limited awareness on the need for male involvement, limited female agency to engage men on ANC, maternal extortion, restrictive gender and socio-cultural norms regarding male ANC attendance, limited engagement of men by ANC staff and intrapersonal fears that fuel the avoidance of antenatal clinics. These overlapped across all three COM-B constructs, and 9 TDF. Overall, we noted that the motivation of male partners to participate in antenatal attendance is strongly influenced by social opportunity factors categorised as restrictive gender, social and cultural norms on male ANC attendance and psychosocial capability underpinned by limited health system engagement and awareness of male role in antenatal care.

**Conclusions:** This study identified multi-dimensional barriers related to male partner capability, opportunity and motivation to participate in antenatal care services. There is a need for interventions that employ gender-transformative approaches to adapt the socio-cultural environment and the messaging on antenatal care for optimal male involvement and subsequently, better health outcomes for mothers and children in Cameroon.

## Introduction

Maternal mortality has remained high despite global efforts to promote safe motherhood as laid out in the Sustainable Development Goals (SDG3.1)(1). Worldwide, 287,000 women died from pregnancy-related complications in 2020(2). Over 99% of these preventable deaths occur in low-income settings with sub-Saharan Africa (SSA) accounting for over 70% of the global burden of maternal deaths(3). Evidence suggests that access to and utilization of quality antenatal care and skilled attendance during pregnancy and childbirth can be an effective strategy to improve maternal and child health (MCH)(4–6). Male involvement in MCH has therefore been proposed as a crucial strategy in resource-limited settings because male partners have economic and decision-making power and consequently significant influence over the health-seeking behaviours of their pregnant partners(7–9). There is currently no agreed definition and indicator for measuring male involvement in MCH. The term varies depending on context—and this ranges from male antenatal care (ANC), immunization or infant welfare attendance, male partner HIV testing during pregnancy, to spousal discussion and men’s domestic and financial support during pregnancy(10–12). In this study, we define male involvement as a man attending ANC with his pregnant partner.

The global recognition of men as key players in MCH has its roots in the 1994 International Conference on Population and Development (ICPD) in Cairo, Egypt(13). The ICPD program of action laid emphasis on male shared responsibility and participation in sexual and reproductive health as a means of achieving gender equality, equity and women’s empowerment(13,14). Research points to the fact that the mechanism through which male involvement impacts maternal and child outcomes is linked to the influence men have over maternal behaviours(7). Furthermore, data from an intervention study across African countries suggests that the three indexes that consistently determine women’s use of antenatal services and skilled birth attendance (SBA) are: a husband’s involvement in maternal decision-making, spousal discussions, and counselling on birth preparedness(15). This has recently been supported by a number of studies that suggest that male involvement in maternal health improves the utilization of prenatal and postnatal services, prevents pregnancy complications and improves overall maternal and infant survival(16–18). Despite these benefits, low levels of male involvement has been reported in SSA with figures as low as 14% in South Africa(19); 26% in Kenya(20); 27.1% in Nigeria(21); and 6-65% in Uganda(22).

Cameroon is currently ranked at the 16^th^ position globally for maternal deaths with an estimated maternal mortality of 438 per 100,000 live births(2). Despite progress in reversing the trend for under-five mortality, the country’s maternal mortality ratio increased from 430 to 782 per 100,000 live births between 1990 and 2011. In addition, the country witnessed a decrease in 1^st^ and 4^th^ antenatal care visits and stagnation in access to skilled personnel during pregnancy between 2004 and 2014. In response to this, the country’s government enacted the 2013 multisectoral program known by its French acronym PNLMNI (Order No. 095 / CAB / PM of 11 November 2013) to reduce maternal and child mortality(23). This was followed by the 2016-2027 Health Sector Strategy that seeks to align the 2013 enactment with the Sustainable Development Goals to target 80% of MCH issues both at the community and health facility levels by 2027(24). Although male involvement is not specifically enshrined in the aforementioned health sector strategy, the National Gender Policy Document (2011–2020) in keeping with the country’s “Vision 2035”, highlights the need for men’s involvement in maternal, reproductive health and HIV/AIDS prevention strategies(25). This has however not been translated to target male involvement in health service delivery models with only 4.7% of men participating in antenatal care with their pregnant partners(26).

Studies in SSA with Cameroon inclusive have reported barriers to male antenatal attendance, including social or normative beliefs that antenatal care is a female affair, lack of time, negative staff attitude and fear of HIV testing(17,27,28). While most of these barriers are reported from the perspectives of women, few studies have reported barriers from the perspectives of men and service providers. Additionally, the majority of the reported studies do not provide the theoretical underpinning and contextual influences on male partner behaviour during the antenatal period. We therefore used the Capability, Opportunity and Motivation (COM-B) Model of Behavioural analysis to contextualise male partner ANC behaviour in order to inform the development of an intervention that is aimed at promoting male involvement in maternal and child health in Cameroon.

The COM-B model posits that factors that influence a given behaviour (B) can be understood through an exploration of how Capability (C), Opportunity (O) and Motivation (M) interact to either enable or hinder the behaviour. This model is central to the Behaviour Change Wheel (BCW)— an encompassing behavioural framework that was developed through the synthesis of 19 behaviour change frameworks in a behavioural theorist meeting that held in the united States of America(29). It has mostly been used in combination with the Theoretical Domains Framework (TDF) which features 14 domains that further expand on COM-B constructs and captures mediating factors on behaviour change(30,31). While COM-B has been applied to design interventions in a variety of contexts like medication adherence, smoking cessation, non-communicable diseases and STI testing, limited evidence exists on its use in developing countries or the design of RMCH interventions(32,33). Regarding male involvement behaviour specifically, there is limited consensus on theoretical approaches that are relevant in the investigation of the determinants of male involvement in Cameroon and sub-Saharan Africa. To address this gap, this study explored barriers to male involvement in antenatal care attendance, in order to provide a theoretical understanding of male involvement behaviour and inform intervention development for pregnant couples in the North West Region of Cameroon.

## Methods

This study was nested in a larger study that is being conducted to develop an intervention to promote male involvement in MCH/HIV during the prenatal period. It represents the formative phase of the study and data collection took place between January and December 2021. We report this study’s methodology and results following guidance from the Consolidated Criteria for Reporting Qualitative Research (COREQ)(34).

### Study design

We chose a qualitative study design underpinned by the naturalistic enquiry approach to explore perspectives on the barriers to male ANC attendance(35). This was suitable as it enabled us to use semi-structured interviews (SSIs) and Focus Group Descriptions (FGDs) to capture participant accounts and provide comprehensive summaries of the factors that impede male participation in antenatal care.

### Study setting

Our study was conducted in Bamenda Health District in the North West Region of Cameroon. The North-West region is a historically disadvantaged, politically unstable and high-density region—currently ranked 3^rd^ most populous in Cameroon with an estimated population of 2 million inhabitants(36). The region has one of the poorest reproductive, maternal and child health indices in the country. Antenatal care coverage of at least one visit was 58% in 2017; childhood immunization coverage was 68% and under five mortality was 57 per 1000 live births(26,37) In 2018, the adult HIV prevalence in the region was estimated at 4.0%, which is higher than the national average while maternal HIV prevalence was estimated at 5.0%(38). The region is largely traditional and patriarchal with prohibitive gender norms and socio-cultural customs that impede male involvement in maternal and child health services(27).

Bamenda Health District is the largest of the 19 districts in the North West Region. The district is located in an urban and peri-urban locality composed of 17 health areas and 35 health facilities—18 public, 12 lay private and 5 confessional serving an estimated population of 800,000 urban and rural residents(36). The main public facility is Bamenda Regional Hospital – a level 2 referral hospital. The district covers a total surface area of 560km^2^ and is centrally located within the city of Bamenda —which serves both as the administrative headquarters of Mezam Division and capital city of the North West Region of Cameroon. The city is cosmopolitan with inhabitants originating across the national territory and neighbouring Nigeria. It is made up of three towns: Mankon, Nkwen and Bamendankwe represented by Bamenda I, II, and III Sub-divisions respectively(36).

Study participants for this study were drawn from Nkwen Baptist Health Centre within Cameroon Baptist Convention Health Services (CBCHS) —a private faith-based NGO in Cameroon. The facility has a modern infrastructure with a 114 bed capacity and 250 staff attending to over 18000 patients monthly(39). It was purposively chosen because it is centrally located within Nkwen town in Bamenda II Sub-division, and has high volume antenatal clinics (over 358 clients per month) with well-established Option B+ services. It also attracts a mix of clients with varying socio-cultural backgrounds that was important for this study

### Participant recruitment and sampling approach

Pregnant or recently postpartum women, male partners and ANC/HIV health workers who were 18 years and above were eligible for inclusion in this study. We employed purposive maximum variation sampling in order to achieve maximal variation regarding age, parity and couple ANC attendance. The variation in our selection was underpinned by the need to capture a wide range of perspectives, perceptions and experiences in order to identify and report common patterns that emerge from heterogeneity(40). We therefore observed group antenatal and post-natal consultations to subjectively identify information-rich participants whom we subsequently approached and invited for interviews or group discussions(41).

We followed recommendations for sample size estimation based on the study question, established evidence on sample size estimates for similar studies and informational redundancy as per the +3 criterion(42,43). A total of 103 eligible women and their male partners were therefore approached of whom 90 consented; four women declined and nine men who were contacted through telephone were unreachable. Of the 80 pregnant women and male partners who accepted our invitation, 44 were women (n= 38 SSIs; n=6, FGDs) and 36 were men (n= 30 SSIs; n=6, FGDs). We purposefully recruited 10 Staff members (two male and eight female) for an FGD based on their clinical roles within ANC/HIV units and their level of education.

To ascertain that saturation was being achieved, we adopted a hybrid approach—data set and individual interviews by discussing the depth and breadth of participant views and perspectives during debriefing sessions, reviewed fieldnotes for recurrent and divergent themes, and noted where infrequent or no new views were expressed(44). Additionally, we probed participant views during interviews until no new information about a particular topic was forthcoming.

### Data collection

The data collection process began with site visits for institutional approval, ANC observation, participant recruitment between January and June 2021. This was closely followed by interviews and FGDs interspaced with debriefing sessions between July and December 2021.

The corresponding author (LHF)— conducted the majority of SSIs (53) and co-facilitated all FGDs. LHF was assisted by a female Bachelor’s degree nurse midwife and a female master’s level sociologist who served as co-interviewers, observers and note-takers during group discussions. The study’s languages were Pidgin (Cameroonian Creole) and English depending on participant preference. We took field notes during interviews and FDGs to capture non-verbal cues and unanticipated events. Although these notes served as an additional source of data, they were not used as primary data sources during analysis. They provided context to participant responses, aid debriefing and enrich analytic memos.

SSIs and group FGDs were conducted using topic guides. The development of these guides was theoretically underpinned by empirical evidence, the National MCH handbook and the COM-B model mapped unto the TDF. Participants were broadly asked: *What are the barriers that prevent men from attending ANC? Why do you perceive them as barriers? How do you feel about these barriers?* We used these guides iteratively with additional prompts beginning with participants’ responses to enrich and add depth to concepts that emerged during interviews and group discussions

With the exception of the staff FGD guide, topic guides and demographic forms were piloted with two postpartum women and one pregnant woman. These were further refined following feedback from participants and the research team. With the exception of two SSIs, all SSIs and FGDs were conducted face-to-face in a private room at the health facility. With permission from participants, interviews and FDGs were audio-recorded, translated and transcribed verbatim. Interviews sessions lasted between 22 – 65 minutes while FGDs lasted between 1-2hrs. Participants were provided snacks and transport reimbursements which ranged between 350frs CFA (40pence) and £1 (750frs CFA).

### Data management and analysis

Both FGDs and SSIs transcripts were anonymized with numbers and combined to represent one data set and managed using NVIVO(45). The first five transcripts were checked against audio for accuracy following the agreed SOP that was developed for this study. The overall approach to analysis was directed content analysis followed by inductive thematic analysis(46,47). Summarily, analysis broadly involved a 7-stage approach with combined guidelines adapted from both Hsieh and Braun as outlined in the supplemental file 1 &2 (46,47). The relevance of themes for inclusion was informed by three criteria: (1) frequency of occurrence, (2) presence of conflicting beliefs (3) perceived strength of the belief to influence the target behaviour(48,49). This was finalized with a tabular representation of themes matched to, belief statements, COM-B constructs and relevant theoretical domains. This matrix (See supplementary file 3) was reviewed by TN and HS for further insight, refinement, interpretation and exploration of dissonant areas.

### Ethical consideration

Ethical approval for this study was granted by the London School of Hygiene and Tropical Medicine ethics committee (Ref: 18003) and the Cameroon Baptist Convention Health Services Internal Review Board (IRB2019-33). Written informed consent was sought from all participants. Participants were assured of confidentiality, anonymity and the non-impact of their participation on the care they were receiving.

## Results

Table 1 presents the socio-demographic characteristics of study participants. Over half (56%) were female; 73% were married, 25% were cohabitating while 3% were single. Our respondents had varied occupations, with 36% in professional employment (excluding staff members), 55% reporting self-employment such as bike or taxi driver, farmer, trader and hair dressing. Majority were literate with (37%) having college or university level education.

**Table 1.** Socio-demographic characteristics of study participants.

| Variable | SSIs (n=68) |  |  |  | Male & Female FGDs (n=12 participants) |  |  |  | Staff FGD (n=10 participants) |  |  |  |
| --- | --- | --- | --- | --- | --- | --- | --- | --- | --- | --- | --- | --- |
|  | Female |  | Male |  | Female |  | Male |  | Female |  | Male |  |
|  | N | % | N | % | N | % | N | % | N | % | N | % |
|  | 38 | 56 | 30 | 44.1 | 6 | 100 | 6 | 100 | 8 | 80 | 2 | 20 |
| **Marital status** |  |  |  |  |  |  |  |  |  |  |  |  |
| Married | 29 | 76 | 20 | 67 | 4 | 66.7 | 5 | 83 | -- | -- | -- | -- |
| Cohabiting | 7 | 18 | 10 | 33 | 2 | 33.3 | 1 | 17 | -- | -- | -- | -- |
| Single | 2 | 5 | -- | -- | -- | -- | -- | -- | -- | -- | -- | -- |
| **Level of Education** |  |  |  |  |  |  |  |  |  |  |  |  |
| Primary or less | 10 | 26 | 7 | 23 | 1 | 16.7 | 2 | 33 | 0 |  | 0 |  |
| Secondary to high school | 13 | 34 | 11 | 37 | 2 | 33.3 | 3 | 50 | 3 |  | 0 |  |
| Completed university | 15 | 40 | 12 | 40 | 3 | 50 | 1 | 17 | 5 |  | 2 |  |
| **Employment status\*** |  |  |  |  |  |  |  |  |  |  |  |  |
| Employed professional | 15 | 40 | 7 | 23 | 3 | 50 | 4 | 67 | -- | -- | -- | -- |
| Self-employed | 16 | 42 | 23 | 77 | 3 | 50 | 2 | 33 | -- | -- | -- | -- |
| Not working | 7 | 18 | --- | --- | -- | -- | -- | -- | -- | -- | -- | -- |
| **ANC Visit with partner\*\*** |  |  |  |  |  |  |  |  |  |  |  |  |
| No | 30 | 83 | 23 | 77 | 3 | 50 | 4 | 67 | -- | -- | -- | -- |
| Yes | 3 | 8 | 7 | 23 | 3 | 50 | 2 | 33 | -- | -- | -- | -- |
| **Mean age** | **31** |  |  |  | **30** |  |  |  | **37** |  |  |  |

\*Not working includes students, housewives and not employed; Self-employed includes bike rider, farmers, traders, hairdresser etc ; \*\* Five missing data on ANC attendance with partner; -- not applicable or data not collected.

Very few (19%) reported that they had attended ANC with their partners. Among staff members, two were male while eight were female among which we had midwives (06), nutritionist(01), an HIV physician (01), an HIV site nurse(01), an Option B+ team lead(01) a PMTCT regional Focal point nurse(01).

Six themes emerged from our analysis: (1) limited awareness/knowledge on the need for male involvement, (2) limited female agency to engage men on ANC, (3) maternal extortion, (4) restrictive gender and socio-cultural norms regarding male ANC attendance, (5) limited engagement of men by ANC staff and (6) intrapersonal fears that fuel the avoidance of ANC clinics.

We conceptually organised these by mapping them to belief statements and theoretical constructs within the capability, opportunity and motivation domains in COM-B model in **Figure1: Conceptual analysis of barriers to male involvement in ANC underpinned by the COM-B model of behaviour change** (29,31)

### Limited awareness on the need for men in antenatal attendance

The majority of participants reported that the low involvement of men in ANC is as a result of the limited awareness and knowledge about their role in MCH and specifically ANC attendance which is generally perceived as a woman’s duty.

> *“I think the very first thing is that men fail to know that they have a part to play during antenatal attendance because they think it is the woman’s duty to come, learn and practice what she has been told”—* Male FGD participant, in their 20s.

> *“He is not really informed about the importance of couple ANC visit as a parent or partner should” —*Female SSI participant, in their 30s.

A subset of study participants who echoed this perspective blamed this lack of awareness on the health system that has not made provision for men by requiring and duly informing them for ANC attendance with their pregnant partners.

> *I blame the ignorance on the part of the health authorities. They have not made a provision for us. If a pregnant woman does not go for antenatal care, even an uneducated grandmother will ask her why because they know it is mandated. However, when a man does not go, nobody will ask him questions because it is not mandated anywhere. I believe that these health institutions should be the ones to make it mandatory and permit men to have their own clinic day or come along with their wives.—* Male FGD participant, in their 30s.

### Limited female agency to initiate male involvement

Based on anecdotal evidence, health providers verbally require pregnant women to bring their partners for ANC attendance. Most of our female participants reported that they do not have the agency to bear the initial responsibility of convincing and involving their male partners for participation in ANC activities. Both men and women in our study therefore expressed the preference for the health system to bear the initial invitation for male involvement—not women who think their partners will not believe them and not a self-initiative from men themselves who don’t wish to intrude because of long-held perceptions that antenatal clinics are spaces reserved for women.

> *“Sometimes I when I tell him that they said men should come for ANC, he thinks I am joking and only want him to be moving around with me. When he sees an invitation from the hospital he would know that it is a serious issue. Otherwise, it will sound like I am the one forcing him to come.”—* Female SSI participant, in their 20s.

### Maternal extortion

Most respondents opined that men generally bear the financial responsibility for prenatal care in Cameroon and some pregnant women prevented their partners from participating in ANC attendance because they preferred to conceal the true cost of accessing ANC services. They did so in the hope of receiving more money than is actually needed. As such, respondents therefore classified these group of women as part of the barriers to male ANC attendance as they fear the presence of male partners could expose their extortionary schemes.

> *“Women are also barriers. They don’t want their partners to come as participant number 5 mentioned because they don’t want the men to know the amount they are spending for ANC. They use pregnancy as a forum to exhort a lot of money from their partners.”*

> *—* Staff FGD participant, in their 40s.

> *Some women don’t also give the opportunity for their husbands to come with them for financial reasons. They don’t want them to know what is happening here and how much is being spent. —* Female SSI participant, in their 30s.

### Restrictive gender, social & cultural norms on male ANC attendance

Apart from the fact that men feel out of place in antenatal clinics, restrictive gender, social and cultural norms on masculinity inhibited the effective participation of men in antenatal care. Gender normative assumptions that men are superior to women and should opt-out of antenatal care in order to maintain their bread-winning role and dominance were some of the factors participants stated for the low participation of men in ANC. Sub-themes that emerged under this are substantiated below with illustrative quotes.

### Male partner sense of superiority about attending ANC

During interviews and group discussions, it was noted that the Cameroonian society is largely patriarchal. Study participants reported that men hold positions of dominance and are considered superior to women to the extent that attending ANC together with other women is a tacit admission that they are in equal standing with women. Respondents therefore intimated that this perception on the superiority of men prevented most men from attending ANC together with their partners. They also noted that some men who eventually made it to ANC clinics decided to sit at the periphery to observe from a non-identifiable distance.

> *“I think there is a cultural [gender] attachment to this. You know, we African men we have a certain way of relating with women—it is a kind of boss-subordinate relationship. I am the head of the family. I have to dictate and the woman follows…Men don’t see themselves and women as being equals. So they don’t feel comfortable sitting and being given health education together with women. A man may feel like, ‘if I go to clinic with my wife, it may appear like my wife and I are equal’” —* Male SSI participant, in their 30s.

### Lack of male social identity in antenatal clinics

Participants reported that men perceive ANC as a woman’s affair because it is largely attended by women who run their own feminine activities which men don’t socially identify with. In the absence of peers or forefathers who modelled the behaviour, the few men who attended ANC felt shy, out-numbered and out of place in a large pool of women who were clapping, singing and dancing to issues that pertain to their pregnancies.

> *The first day I went, I was in a pool of women and all eyes were on me. I was like…OK “what am I doing here?” I am not pregnant! —*Male SSI participant, in their 30s.

> *“I am always scared of going for ANC. Imagine being the only man among hundreds of women and the stares you will receive. It makes me feel a type…there are songs that women will be singing, clapping and even dancing to… and they may be expecting you to be clapping and singing as well [laughs]. When you are not clapping because you don’t identify, it becomes a call for concern. When you sing along for solidarity, it is still a call for concern because it may appear as though you are pregnant as well, of which you are not!”*

> *—*Male SSI participant, in their 30s.

### Engendered perception of the time value of ANC attendance

The notion that time is not created equally for men and women in Cameroon emerged as an important factor in this study. Participants reported that the majority of Cameroonian men were primary bread-winners—and being able to provide was a source of masculine identity and pride. This came with the expectation for men to appear busy in income-generating activities. Respondents did not therefore rate ANC attendance as an income-generating activity. They rather perceived it as an opportunity cost—a liability to their bread-winning. Respondents also acknowledged that while ANC attendance was an obligatory activity for women it was often perceived as an optional affair for men who did not wish to spend long hours in ANC clinics.

> *“The point is that coming to the clinic for me is a must. Time or no time I must be at the clinic on the appointed date until delivery. That is not the same for my husband. As a man, he must go out and struggle to work so that he can get something (money) to give me for ANC attendance.” —*Female SSI participant, in their 30s.

> *“You have the issue of time…time factor which is also something so important because the man is always busy from morning till night… and remember that it is the man who in most cases (I can say about 80% or 70% of cases), the man is the one who provides everything in the house. So, he makes sure that he catches up with those activities in order to be able to provide. Because when he sits somewhere and loses a day, it is really something big that he has lost” —*Male SSI participant, in their 30s.

### “Woman Wrapper” stigma—male fear of losing control and female fear of appearing in control

Most Cameroonian men from the North West Region have been conditioned to exercise dominance over women. Since ANC has traditionally been an activity reserved for women, the introduction of men into this space was perceived by study participants as an attempt to usurp their power and relegate men to the background. The mental picture that the majority of participants presented about male ANC attendance was a woman at the forefront and the man behind her as a follower. Men who therefore made attempts to attend ANC clinics were mocked by community members and ridiculed as weaklings who have given up their masculine power or are under metaphysical forces from women.

According to several participants, some Cameroonian phrases or idioms have been coined to label men who are in favour of accompanying their partners for ANC. ‘Woman wrapper’ which literally translates to being too attached and subdued under a woman’s loin cloth was one of such derogatory terms. According to participants, this describes a man who takes orders from his wife or always follows her around.

> *“To add to what my brother has said, let me speak from my community. It appears like, attending ANC with a woman is a way of giving up your authority as a man. When your fellow men see you, they look at you like you are a woman [group laughter]. Yes, you are a woman and not the man because if you were a man, you will not be following your wife to go do women’s things” —*Male FGD participant, in their 30s.

The need for men to maintain dominance is so normative in the Cameroonian context to the extent that even women who attended ANC with their partners reflectively defended their partner’s choice in order to distance themselves from any perceived notion that they are controlling their husbands.

> *“ You know, African men have a mentality that if they follow their wives for antenatal care, people might see them and think that he lacks something to do or he is a “woman wrapper” [weakling, sissy or subdued man]…The day we came for ANC, he was the only man who came for ANC and even though some women will think that I am controlling my husband, that was his personal decision” —* Female SSI participant, in their 20s.

### Limited engagement of men by ANC staff

Health system factors were also thought to limit male participation in antenatal care. Staff and participants who had previously attended ANC opined that the engagement of men by ANC staff was ridden with tokenism which some men find derogatory. Others felt their presence in ANC made no difference because the focus was mostly on their pregnant partners. Sub-themes that emerged from this are explained below:

### Token-based engagement (ANC claps for men who attend)

Participants reported that men who were spotted in ANC clinics were given a special ‘ANC clap’ as a form of recognition. While this was generally good-intentioned on the side of service providers, some men received this with mixed feelings. ‘ANC claps’ for men who attended clinics was perceived as an insult to their identity. They felt it reduced them to preschool children who have to be clapped for, for fulfilling the bare minimum of ANC attendance with their wives—something they do not have to be coerced with claps to do.

> *“Some men have said: “I came to the clinic and at the end they said ‘let’s clap for papa, papa came for clinic today’ and they felt like they were in primary school. So they will not come again, because they don’t like being treated as kids” —*Staff FGD participant in their 30s.

### Lack of male engagement and female-focused health education

Some participants reported that health facilities have not made provision for their presence in ANC clinics. This is evident in the fact that male attendance has not been required and for those who made the effort to attend, providers rarely engaged them. Rather, the entire focus was on their pregnant partners. Some participants substantiated this with the fact that the need for couple attendance is something they only heard about through this study.

> *“You are the first to bring up this procedure on couple ANC attendance and testing for HIV. That is what we should have done before but the attention of the hospital has mostly been on her.” —*Male SSI participant, in their 40s.

### Intrapersonal Emotions (Fear)

Study participants across FGDs and SSIs echoed the fact that male ANC attendance evokes a range of emotions that serve as barriers to their attendance, including fear. Sub-themes below further expand on these:

### Fear of being judged or discovered for extra-marital affairs

Participants mentioned that a pregnant woman in the Cameroonian context is more of an abandoned sexual project and most men use this period to visit their “deuxième bereau”— local lexicon denoting experimentation with extra-marital relationships. They [men] therefore avoided ANC clinics because attending might expose their deeds. A minority of participants equally noted that some aspects of health education during ANC have a judgmental tone that might prick the consciences of men involved in such practices.

> *“Once a woman becomes pregnant, some men get into the practice of what we call ‘side chicks’… they avoid ANC all-together because they don’t want to be judged about their sinful lives and extra-marital affairs. They don’t want health talks at the clinic to echo in their mind and their consciences*” —Female SSI participant, in their 30s.

### Fear of HIV testing

Male involvement in the Cameroonian context is largely driven by the HIV epidemic and need for PMTCT to the extent that participants associated male ANC attendance with HIV testing. As such, participants reported that men feared attending ANC because they will be required to test for HIV if they came with their pregnant partners.

> *“The ANC testing requirement is also a barrier. Perhaps they are aware that if they come, they will be checked and tested and for those who are unaware/unsure of their status, they don’t want to come” —* Male SSI participant, in their 30s.

> *“If a woman is coming for ANC and she tells her husband that they are going to do HIV testing for both of them, at that point you will hear the man say ‘my coming is not necessary’. The name HIV alone cancels the whole issue” —*Female SSI participant, in their 20s.

### Avoidance of responsibility

In a setting like Cameroon where paternity is rarely established through civil and legal means, male ANC attendance is seen as an explicit form of taking both paternal and financial responsibility and some men want to shy away from this. Additionally, the wide practice of extra-marital affairs during pregnancy probably resulted in pregnancies that most men did not want to be publicly associated with through ANC attendance.

> *“I think there is also a financial barrier. Some men don’t want to attend ANC because attending will mean they are taking responsibility as authors of the pregnancy and this also means they are required to show up as fathers and engage in financial responsibilities”*

> *—*Male SSI participant, in their 30s.

> *“Some men are ashamed especially when they have impregnated many girls in the neighbourhood and they don’t want to be tagged as a particular woman’s husband when there are other women he has impregnated and he is denying being responsibility for their pregnancies.” —*Female SSI participant, in their 20s.

## Discussion

This research paper sought to identify and characterize barriers to male partner involvement in antenatal care in the North West Region of Cameroon. Our study findings illustrate the complexity of factors that influence men’s perceptions and behaviour towards ANC attendance. We conceptualised these using the COM-B model of behaviour and TDF with the main barriers reported as: limited awareness on the male role in ANC, inadequate health system engagement, fear of judgement and HIV testing, and restrictive gender and social norms on male ANC attendance.

The COM-B component of Opportunity was highly salient in this study, with social opportunity strongly representing the viewpoints of participants through four thematized barriers: male superiority, lack of male social identity in ANC clinics, engendered perception of time and fear of the ‘woman wrapper’ stigma. Significant among these is the widespread belief that antenatal attendance is a female affair and men who engage in antenatal-seeking behaviours are seen as jobless, “weak” and under the control of their wives. As a result, the participation of men in ANC activities is socially outlawed as it competes with their bread-winning roles and shapes perception on the identity, masculine credibility and engagement of men who venture into antenatal clinics. These perceptions on male ANC attendance has been echoed across previous studies in sub-Saharan Africa(8,28,50,51). Our study further highlights the engendered dimension on male social identity and the ‘woman wrapper’ stigma associated with men who accompany their partners for ANC. This underscores the need to reconstruct ANC services through a theoretical and gender-transformative approach that strategically considers prevailing socio-cultural and gender norms in male involvement programming in Cameroon.

Physical opportunity took the form of the organizational culture where the health system does not adequately initiate male involvement and fails to engage male partners when they make the effort to attend ANC alongside their pregnant partners. For the few men who had experienced ANC attendance the ‘ANC clap’, which seeks to recognise their presence, was perceived as a reinforcement of their exclusion. Our female participants equally reported that they do not have the agency to bear the responsibility of involving their partners for antenatal care. This stems from the current health system practice which requires pregnant women to hand invitation letters to their partners or verbally inform them on the need for couple ANC attendance –a common practice as reported by similar studies in Sub-Saharan Africa(52). This finding highlights that the lack of male involvement awareness limits men’s perceptions of psychological capability and informed decision-making on the need to attend ANC with their partners.

Further analysis on the health system reliance on women as relay agents to men revealed that information on male involvement from women is perceived by men as incomplete or exaggerated. As such, participants in our study proposed that the health system needs to go beyond the traditional *‘go and bring your husband’* agenda by extending a direct appeal to men through health and gender sensitive messaging. While this recommendation is consistent with recent study implications in Malawi and Zambia(53), it is in stark contrast to a study in Tanzania in where participants proposed that women should bear the emotional and intellectual burden of involving their partners in maternal and child health(54).

Psychological capability was equally a key domain, which underpins male involvement in ANC through limited awareness and knowledge on the need for male partner participation in antenatal activities. Consistent with our study, evidence from studies in South Africa and Ghana demonstrated that the lack of awareness on the need for male involvement influenced male behaviour and non-participation in ANC activities(55,56). Despite the documentation of high levels of knowledge regarding ANC activities by Nkuoh and colleagues in Cameroon(27), our study demonstrates that this does not necessarily translate to interest in male involvement. This disconnect between knowledge and interest could likely be as a result of the perception that ‘clinic’ as referenced by participants is a female-focused interventions. To counter this, we therefore argue for the health system to tailor communication on antenatal education to messaging that engages men directly, reflects need and the inclusivity of men in the antenatal care package.

Further to inadequate health system engagement and motivation, participants in our study associated the involvement of men in antenatal care with HIV testing. This might be due to the fact that the initiation of male involvement programs in Cameroon and other African settings historically focused on their role in PMTCT(27,28,57). Additionally, the fear men expressed for HIV testing and its possible outcome in antenatal settings could be attributed to the prevailing gender norm that men are strong and should therefore not be sick or seen in spaces generally reserved for weak and vulnerable members of the society like women and children. It is against this backdrop that male ANC attendance is highly stigmatised. Thus, the need to go beyond applauding mere ANC attendance or HIV testing to the involvement of men in clinical assessments like foetal heart monitoring, health education and convenient ANC scheduling. These could potentially reframe the stigmatization of male ANC attendance and impact motivation to participate in ANC activities.

This is the first study in Cameroon to identify and conceptualise factors that influence male engagement in ANC. A unique feature in our study is the use of the COM-B model and TDF as an additional step following thematic analysis to characterise male involvement behaviour around ANC. The endorsement of barriers across COM-B components demonstrates the complexity of male involvement behaviour in the North West Region of Cameroon. With COM-B centrally located within the behaviour change wheel, the identified factors in our study can be linked to appropriate intervention functions and behaviour change techniques that could address barriers to male involvement in ANC

Our study should however be interpreted in light of methodological and practical constraints that limit the generalizability of our findings. First, specific gender and socio-cultural norms that characterise the North West Region may limit the extent to which our results are generalizable to other regions in Cameroon. Notwithstanding, there was no indication that our findings on the barriers to male involvement in ANC differ significantly from the prevailing literature in Cameroon and Africa at large.

## Conclusion

Our study drew upon the diverse perspective of pregnant women, male partners and health workers to explore the barriers to male ANC attendance in the North West Region of Cameroon. We found that the low motivation of male partners to participate in antenatal care is at the intersection of social and physical opportunities (socio-cultural and gender norms) and psychological capability (limited knowledge and agency). Based on this finding, we recommend that the development of interventions for male involvement in a patriarchal setting like the North West Region of Cameroon should target the low levels of awareness and direct engagement of male partners with messaging that override restrictive gender norms.

## Supporting information

Code book

Data analysis

Coding matrix

Conceptual analysis

## Data Availability

All relevant data used to reach the conclusions drawn in the study are within the paper and its Supporting Information files. This data has also been safely stored at the CBCHS- https://cbchealthservices.org/ and LSHTM-https://www.lshtm.ac.uk. For reasons of confidentiality, original transcripts and demographic information is not available to the public as some of the interviewees are easily identifiable. This data could however be made available upon reasonable request.

## Acknowledgments

The authors would like to thank the Cameroon Baptist Convention Health Services for granting us permission to work within their facility. We acknowledge Ms Nancy Nyakieh (NN), for assisting with interviews and preliminary analysis, Mrs Tiyang Monica (TM) for assisting with data collection—Focus Group observation and note-taking and Mrs Tibah Tchouba Carine-Flore for her transcription and translation services. Finally, we wish to extend our gratitude to all participants who participated in this study.

## Supporting Information

S1 Text. Code Book

S2 Text. Data Analysis stages

S3 Text. Coding Matrix

S4_Fig.PDF Conceptual Analysis of Barriers

## References

1. UNDESA. Transforming our world: the 2030 Agenda for Sustainable Development | Department of Economic and Social Affairs [Internet]. 2015 [cited 2023 Sep 6]. Available from: https://sdgs.un.org/2030agenda

2. WHO. Trends in maternal mortality 2000 to 2020 estimates by WHO, UNICEF, UNFPA, World Bank Group and UNDESA/Population Division. Geneva; 2020.

3. WHO. Maternal mortality, Key Facts. [Internet]. 2023 [cited 2023 Aug 14]. Available from: https://www.who.int/news-room/fact-sheets/detail/maternal-mortality

4. Campbell OM, Graham WJ. Strategies for reducing maternal mortality: getting on with what works. Lancet [Internet]. 2006 Oct 7 [cited 2023 Aug 5];368(9543):1284–99. Available from: http://www.thelancet.com/article/S0140673606693811/fulltext

5. Brown CA, Sohani SB, Khan K, Lilford R, Mukhwana W. Antenatal care and perinatal outcomes in Kwale district, Kenya. BMC Pregnancy Childbirth [Internet]. 2008 Jan 10 [cited 2023 Aug 5];8(1):1–11. Available from: https://bmcpregnancychildbirth.biomedcentral.com/articles/10.1186/1471-2393-8-2

6. Carroli G, Villar J, Piaggio G, Khan-Neelofur D, Gülmezoglu M, Mugford M, et al. WHO systematic review of randomised controlled trials of routine antenatal care. Lancet [Internet]. 2001 May 19 [cited 2023 Aug 14];357(9268):1565–70. Available from: https://pubmed.ncbi.nlm.nih.gov/11377643/

7. Ngom P, Debpuur C, Akweongo P, Adongo P, Binka FN. Gate-keeping and women’s health seeking behaviour in Navrongo, northern Ghana. Afr J Reprod Health. 2003;7(1):17–26.

8. Kwambai TK, Dellicour S, Desai M, Ameh CA, Person B, Achieng F, et al. Perspectives of men on antenatal and delivery care service utilisation in rural western Kenya: a qualitative study. BMC Pregnancy Childbirth [Internet]. 2013 Jun 21 [cited 2023 Jul 23];13:134. Available from: /pmc/articles/PMC3691751/

9. World Health Organisation. WHO recommendations on health promotion interventions for maternal and newborn health. Geneva: WHO; 2015.

10. Mkandawire E, Hendriks SL. A qualitative analysis of men’s involvement in maternal and child health as a policy intervention in rural Central Malawi. BMC Pregnancy Childbirth [Internet]. 2018 Jan 19 [cited 2023 Dec 18];18(1):1–12. Available from: https://bmcpregnancychildbirth.biomedcentral.com/articles/10.1186/s12884-018-1669-5

11. Ditekemena J, Koole O, Engmann C, Matendo R, Tshefu A, Ryder R, et al. Determinants of male involvement in maternal and child health services in sub-Saharan Africa: A review. Reprod Health [Internet]. 2012 Nov 21 [cited 2023 Oct 19];9(1):1–8. Available from: https://reproductive-health-journal.biomedcentral.com/articles/10.1186/1742-4755-9-32

12. Alio AP, Lewis CA, Scarborough K, Harris K, Fiscella K. A community perspective on the role of fathers during pregnancy: A qualitative study. BMC Pregnancy Childbirth [Internet]. 2013 Mar 7 [cited 2023 Nov 16];13(1):1–11. Available from: https://bmcpregnancychildbirth.biomedcentral.com/articles/10.1186/1471-2393-13-60

13. EuroPROFEM. Male Involvement in Reproductive Health: ICPD-Family Care International [Internet]. [cited 2023 Apr 11]. Available from: http://www.europrofem.org/contri/2_04_en/en-masc/53en_mas.htm

14. Lazarus JV. The changing role of men since ICPD [Internet]. Entre Nous. Copehagen, Denmark: Entre Nous Cph Den ; 1999 Apr [cited 2023 Apr 11]. Available from: https://pubmed.ncbi.nlm.nih.gov/12222325/

15. Mwakayongwe T, Brazier E, Mutunga A, Perkins M, Themmen E, Wa U, et al. Testing Approaches for Increasing Skilled Care During Childbirth: Key Findings [Internet]. 2007 [cited 2023 Aug 14]. Available from: http://www.childinfo.org/areas/deliverycare/countrydata.php

16. Tokhi M, Comrie-Thomson L, Davis J, Portela A, Chersich M, Luchters S. Involving men to improve maternal and newborn health: A systematic review of the effectiveness of interventions. PLoS One [Internet]. 2018 Jan 1 [cited 2023 Apr 21];13(1):e0191620. Available from: https://journals.plos.org/plosone/article?id=10.1371/journal.pone.0191620

17. Yargawa J, Leonardi-Bee J. Male involvement and maternal health outcomes: systematic review and meta-analysis. J Epidemiol Community Health [Internet]. 2015 Jun 1 [cited 2023 Apr 13];69(6):604–12. Available from: https://jech.bmj.com/content/69/6/604

18. Lundgren RI, Gribble JN, Greene ME, Emrick GE, De Monroy M. Cultivating Men’s Interest in Family Planning in Rural El Salvador. Stud Fam Plann [Internet]. 2005 Sep 1 [cited 2023 Apr 21];36(3):173-88. Available from: https://onlinelibrary.wiley.com/doi/full/10.1111/j.1728-4465.2005.00060.x

19. Yende N, Van Rie A, West NS, Bassett J, Schwartz SR. Acceptability and Preferences among Men and Women for Male Involvement in Antenatal Care. J Pregnancy. 2017;2017.

20. Aluisio AR, Bosire R, Bourke B, Gatuguta A, Kiarie JN, Nduati R, et al. Male Partner Participation in Antenatal Clinic Services is Associated with Improved HIV-free survival Among Infants in Nairobi, Kenya: A Prospective Cohort Study. J Acquir Immune Defic Syndr [Internet]. 2016 Oct 10 [cited 2023 Jul 17];73(2):169. Available from: /pmc/articles/PMC5023460/

21. I. OBA, O. AOE, O. AA, A. AA, O. OS. Perception, attitude and involvement of men in maternal health care in a Nigerian community. J Public Health Epidemiol [Internet]. 2013 Jun 30 [cited 2023 Jul 17];5(6):262–70. Available from: https://academicjournals.org/journal/JPHE/article-abstract/7BF1FF25254

22. Francis Kariuki K, Seruwagi GK, Yu R. Determinants of Male Partner Involvement in Antenatal Care in Wakiso District, Uganda. J Adv Med Med Res [Internet]. 2016 Nov 5 [cited 2023 Jul 17];18(7):1–15. Available from: https://journaljammr.com/index.php/JAMMR/article/view/254

23. Minsante (MOH). National Multisector Program to Combat Maternal, Newborn and Child Mortality (PLMI-CMR) [Internet]. 095 / CAB / PM Ministry of Health, Cameroon; Nov 13, 2013. Available from: https://www.plmi.cm/

24. Ministry of Public Health (MINSANTE). Health Sector Strategy 2016 - 2027 | [Internet]. Yaounde, Cameroon; 2017 [cited 2023 Apr 17]. Available from: https://www.minsante.cm/site/?q=en/content/health-sector-strategy-2016-2027-0

25. Japan International Cooperation Agency (JICA). 2015 Country Report of Gender Profile (Cameroon). 2015.

26. NACC. PMTCT Progress Report 2017 [Internet]. Available from: http://www.cnls.cm/sites/default/files/rapport_progres_ptme_ndeg_12_2017_1.pdf

27. Nkuoh GN, Meyer DJ, Tih PM, Nkfusai J. Barriers to Men’s Participation in Antenatal and Prevention of Mother-to-Child HIV Transmission Care in Cameroon, Africa. J Midwifery Womens Health. 2010;

28. Morfaw F, Mbuagbaw L, Thabane L, Rodrigues C, Wunderlich AP, Nana P, et al. Male involvement in prevention programs of mother to child transmission of HIV: a systematic review to identify barriers and facilitators. Systematic reviews. 2013.

29. Michie S, van Stralen MM, West R. The behaviour change wheel: A new method for characterising and designing behaviour change interventions. Implementation Science. 2011;

30. Cane J, O’Connor D, Michie S. Validation of the theoretical domains framework for use in behaviour change and implementation research. Implementation Science. 2012 Apr 24;7(1).

31. Michie S, Atkins L, West R. The Behaviour Change Wheel: A Guide to Designing Interventions. The Behavior Change Wheel: Book Launch Event. 2014. 1–46 p.

32. Han S, Middleton PF, Bubner TK, Crowther CA. Women’s views on their diagnosis and management for borderline gestational diabetes mellitus. J Diabetes Res. 2015;

33. Fulton E, Brown K, Kwah K, Wild S. StopApp: Using the Behaviour Change Wheel to Develop an App to Increase Uptake and Attendance at NHS Stop Smoking Services. Healthcare. 2016 Jun 8;4(2):31.

34. Tong A, Sainsbury P, Craig J. Consolidated criteria for reporting qualitative research (COREQ): a 32-item checklist for interviews and focus groups. Int J Qual Health Care [Internet]. 2007 Dec [cited 2022 Mar 29];19(6):349–57. Available from: https://pubmed.ncbi.nlm.nih.gov/17872937/

35. Sandelowski M. Focus on research methods: Whatever happened to qualitative description? Res Nurs Health. 2000;23(4):334–40.

36. Communication Service at the Governor’s Office. North-West Region’s Web Portal [Internet]. [cited 2022 Feb 25]. Available from: http://www.northwest-cameroon.com/home-35-front-0.html

37. The DHS Program - Cameroon: DHS, 2018 - Cameroon 2018 Demographic and Health Survey - Summary Report (English) [Internet]. 2018 [cited 2023 Apr 21]. Available from: https://dhsprogram.com/publications/publication-sr266-summary-reports-key-findings.cfm

38. Ministry of Health. Cameroon Population-based HIV Impact Assessment (CAMPHIA) 2017-2018: Final Report. . Yaounde; 2020 Dec.

39. Cameroon Baptist Convention Health Services. Nkwen Baptist Health Center-Overview [Internet]. 2019 [cited 2022 Feb 25]. Available from: https://cbchealthservices.org/health-centers/north-west-region/nkwen-baptist-hc/

40. Palinkas LA, Horwitz SM, Green CA, Wisdom JP, Duan N, Hoagwood K. Purposeful sampling for qualitative data collection and analysis in mixed method implementation research. Adm Policy Ment Health. 2015 Sep 22;42(5):533.

41. Creswell JW, Plano Clark VL. Designing and Conducting Mixed Methods Research. International Student Edition. 2018.

42. Marshall Bryan, Cardon Peter, Poddar Amit, Fontenot Renee. Does sample size matterin qualitative research? A review of qualitative interviews in is research. Journal of computer information systems. 2013.

43. Francis JJ, Johnston M, Robertson C, Glidewell L, Entwistle V, Eccles MP, et al. What is an adequate sample size? Operationalising data saturation for theory-based interview studies. Vol. 25, Psychology & Health. 2010.

44. Saunders B, Sim J, Kingstone T, Baker S, Waterfield J, Bartlam B, et al. Saturation in qualitative research: exploring its conceptualization and operationalization. Qual Quant. 2018 Jul 1;52(4):1893.

45. NVivo - Lumivero [Internet]. [cited 2023 Apr 24]. Available from: https://lumivero.com/products/nvivo/

46. Hsieh HF, Shannon SE. Three approaches to qualitative content analysis. Qual Health Res. 2005 Nov;15(9):1277–88.

47. Braun V, Clarke V. Using thematic analysis in psychology. Qual Res Psychol. 2006;3(2):77–101.

48. Francis JJ, Stockton C, Eccles MP, Johnston M, Cuthbertson BH, Grimshaw JM, et al. Evidence-based selection of theories for designing behaviour change interventions: Using methods based on theoretical construct domains to understand clinicians’ blood transfusion behaviour. Br J Health Psychol. 2009 Nov;14(4):625–46.

49. Gale NK, Heath G, Cameron E, Rashid S, Redwood S. Using the framework method for the analysis of qualitative data in multi-disciplinary health research. BMC Med Res Methodol [Internet]. 2013 Sep 18 [cited 2023 Mar 30];13(1):1–8. Available from: https://bmcmedresmethodol.biomedcentral.com/articles/10.1186/1471-2288-13-117

50. Greene ME, Barker G. Masculinity and Its Public Health Implications for Sexual and Reproductive Health and HIV Prevention.

51. Craymah JP, Oppong RK, Tuoyire DA. Male Involvement in Maternal Health Care at Anomabo, Central Region, Ghana. Int J Reprod Med. 2017;2017:1–8.

52. Takah NF, Kennedy ITR, Johnman C. The impact of approaches in improving male partner involvement in the prevention of mother-to-child transmission of HIV on the uptake of maternal antiretroviral therapy among HIV-seropositive pregnant women in sub-Saharan Africa: A systematic review and meta-analysis. BMJ Open. 2017.

53. Mweemba O, Zimba C, Chi BH, Chibwe KF, Dunda W, Freeborn K, et al. Contextualising men’s role and participation in PMTCT programmes in Malawi and Zambia: A hegemonic masculinity perspective. Glob Public Health [Internet]. 2022 [cited 2023 Jul 26];17(9):2081–94. Available from: 10.1080/17441692.2021.1964559

54. Maluka S, Japhet P, Fitzgerald S, Begum K, Alexander M, Kamuzora P. Original research: Leaving no one behind: using action research to promote male involvement in maternal and child health in Iringa region, Tanzania. BMJ Open [Internet]. 2020 Nov 14 [cited 2023 Jul 26];10(11). Available from: /pmc/articles/PMC7668372/

55. Ganle JK, Dery I. ‘What men don’t know can hurt women’s health’: a qualitative study of the barriers to and opportunities for men’s involvement in maternal healthcare in Ghana. Reprod Health [Internet]. 2015 Oct 10 [cited 2023 Jul 25];12(1):1–13. Available from: https://reproductive-health-journal.biomedcentral.com/articles/10.1186/s12978-015-0083-y

56. Mohlala BKF, Gregson S, Boily MC. Barriers to involvement of men in ANC and VCT in Khayelitsha, South Africa. 101080/095401212012668166 [Internet]. 2012 Aug 1 [cited 2023 Jul 25];24(8):972–7. Available from: https://www.tandfonline.com/doi/abs/10.1080/09540121.2012.668166

57. Manjate Cuco RM, Munguambe K, Osman NB, Degomme O, Temmerman M, Sidat MM. Male partners’ involvement in prevention of mother-to-child HIV transmission in sub-Saharan Africa: A systematic review. Sahara J. 2015;12(1):87–105.

