## Supplementary material for "“Go and bring your husband”: a COM-B guided qualitative study on the barriers to male involvement in antenatal care in Bamenda Health District, Cameroon": Code book

**COM-B/TDF Guided Codebook on Barriers to Male ANC Attendance** ^45^

| **Relevant TDF code & definition*** | **TDF Construct** | **COM-B Category** | **Adapted TDF definition**  **(Apply this code to data if it mentions:)** |
| --- | --- | --- | --- |
| 1. **Knowledge**   An awareness of the existence of something | Knowledge of service/condition or scientific rationale | Capability-Psy | Knowledge/awareness on male role in ANC attendance or affirms role upon prompt on need for male ANC attendance |
| 2. **Skills**   An ability or proficiency acquired through practice | Interpersonal skills | Capability-Phy | - Skill required to perform or facilitate male ANC attendance - Participant struggles to communicate need for interpersonal couple ANC attendance or need for providers to take leadership on engaging male partners for couple ANC attendance |
| 3. **Social/Professional Role and Identity**   A coherent set of behaviours and displayed personal qualities of an individual in a social or work setting | Social Identity/Identity | Opportunity | Views on how men perceive their roles in pregnancy or mention their ANC attendance experience |
| 4. **Beliefs about Capabilities**   Acceptance of the truth, reality, or validity about an ability, talent, or facility that a person can put to constructive use | Self-esteem | Capability-Psy | Any complex (superior or inferior) men feel about attending ANC with their partners. Mention of ability to attend ANC with partner |
| 5. **Optimism**   The confidence that things will happen for the best or that desired goals will be attained | Optimism and Pessimism | Motivation | Anticipated benefits or outcome for couple ANC attendance |
| 6. **Beliefs about Consequences**   Acceptance of the truth, reality, or validity about outcomes of a behaviour in a given situation | Outcome expectancies/ consequents | Motivation | Anticipated benefits or outcome of male ANC attendance or non-attendance |
| 7. **Reinforcement**   Increasing the probability of a response by arranging a dependent relationship, or contingency, between the response and a given stimulus | Reward | Motivation | Perceived outcomes of attending ANC like fast-tracking of services for women who attend with their partners or access to expert knowledge |
| **11. Environmental Context and Resources**  Any circumstance of a person's situation or environment that discourages or encourages the development of skills and abilities, independence, social competence, and adaptive behaviour | Organisational culture and climate  Resources | Opportunity-Phy | Issues in the social service policy, hospital environment, clinical triage or resources ( time & money) that (can) influence the decision for male ANC attendance. |
| **12. Social influences**  Those interpersonal processes that can cause individuals to change their thoughts, feelings, or behaviours | Social norms  Modelling  Social comparisons | Opportunity | **Do participants find male ANC attendance acceptable in the Cameroonian context?** Cultural and social norms that influence male engagement in antenatal attendance   - Formal rules that discourage the behaviour |
| **13. Emotion**  A complex reaction pattern, involving experiential, behavioural, and physiological elements, by which the individual attempts to deal with a personally significant matter or event | Positive or negative effect emotions  Fear | Motivation | Positive or negative emotions participants feel about male antenatal attendance or fears that influence the behaviour |

*Out of the 14 TDF #7, 8, 9, 10 & 14 were not found to be relevant to this study’s research questions.
