## Supplementary material for "“Go and bring your husband”: a COM-B guided qualitative study on the barriers to male involvement in antenatal care in Bamenda Health District, Cameroon": Data analysis

**Table 1: Directed Content and Thematic Analysis adapted from Hsieh and Braun**^48,49^

| **Stages** | **Explanation** |
| --- | --- |
| 1. **Data Familiarisation** | Repeated reading and annotation of transcripts for immersion to gain an intimate knowledge of the data. While reading, key ideas and broad analytic thoughts were generated for later review. |
| 2. **Deductive Coding** | Descriptive labelling of a subset of the data (15 purposively selected transcripts) with succinct tags/codes to identify attributes that were relevant to COM-B, TDF and research questions. |
| 3. **Development of Coding framework** | Categorization or grouping of initial codes to generate contextual descriptions and inferential meanings for later application on the entire dataset. This ended with the development of a COM-B/TDF guided codebook (See supplementary file 1) that was later applied to the remaining data set for thematic analysis. |
| 4. **Theme Identification** | Inductive application of coding framework on the entire dataset to generate candidate themes and belief statements that represent tentative themes. |
| 5. **Theme Review** | Testing candidate themes against the entire dataset for coherency and consistency in the theme narrative. |
| 6. **Theme Refinement** | Amalgamation and linking of candidate themes into dominant categories. This involved detailed analysis of each theme to decide its name, scope and whether or not it needs to be split or left to stand alone. |
| 7. **Interpretation and Writing up** | Description of themes, reflection and explanation of patterns within themes and reporting with illustrative participant quotes that back up themes. |
