## Supplementary material for "“Go and bring your husband”: a COM-B guided qualitative study on the barriers to male involvement in antenatal care in Bamenda Health District, Cameroon": Coding matrix

**Barriers to Male Antenatal Attendance: Salient themes Mapped to COM-B and TDF**

| **BARRIERS** | | | | |
| --- | --- | --- | --- | --- |
| **Themes and Sub themes** | **Belief Statements** | **COM-B** | **TDF** | **Illustrative Quotes** |
| 1. **Limited awareness on the need for male antenatal attendance** | There is low awareness on the need for male ANC attendance in Cameroon.  Men are not adequately engaged with information on couple ANC attendance. As such, they don’t see the need for it. | Capability | Knowledge | “I think the very first thing is that men, some men fail to know that they have a part to play during antenatal because it is the woman’s duty to come, learn and practice what she has been told” Male FGD #5  “Those are the two main reasons It’s either they are too busy or they are not aware that they are supposed to come. They assume that ANC is for the woman and the child and they have no part to play. Perhaps they know they just have to give them the money to go ahead” Male #45  “He’s not really informed about the importance of couple ANC visit as parents should” Female #16  “Just to add to that, I think one of the barriers here is ignorance because here we are talking about partners, I mean male partners and who else knows male partner more than the pregnant woman. So, if we do not educate the women themselves to know why it is even important that their partner’s HIV status is well established, then they might become apathetical and indifferent to whether or not their partners come for ANC and know their status”  Male FGD #1 |
| 2. **Limited female agency to initiate male involvement** | Pregnant women don’t feel empowered enough to bear the initial responsibility for partner involvement in ANC attendance.  Health care providers need to take the lead in reaching out to male partners about the need for couple ANC attendance  Women don’t convey antenatal messaging or the need for male involvement properly and this deters men from taking them serious | Capability | Interpersonal skills | “Sometimes I when I say they said that men should come for ANC, he will think I am joking and only want him to be moving around with me. When he sees an invitation from the hospital he would know that it is a serious issue. Let it not sound like I am the one forcing him to come.” Female #1  “Sometimes you listen to lectures and when you share with him, he thinks you are lying” Female #6  “It is only through the hospital’s invitation because there are some men that will not take ANC attendance seriously if it comes from their wives but if they see something directly from the doctor they will know it is serious and will want to come Female #22  “I believe that accompanying my wife for ANC is part of my responsibility but that invitation to participate should come from the ANC clinic. I don’t think I just have to impose myself and be among women because I have my own perspective about things. I just believe I should be invited. I don’t like intruding into people…maybe people have their own protocols they follow” Male #44 |
| 3. **Perceptions on male partner identity in antenatal spaces**  - Lack of male social identity in antenatal clinics - Men feel out of place in ANC clinics | Men perceive ANC as a woman’s thing since they are not the carriers of the pregnancy.  ANC clinics is not the place for men. ANC is for women’s affairs  The large number of pregnant women in antenatal spaces is over-powering to men  Couple ANC attendance has not been normalised and the low turnout of men who accompany their pregnant partners for ANC is not encouraging to other men | Opportunity | Identity/Social identity | “I am always scared of going for ANC. Imagine being the only man among hundreds of women and the stares you will receive. It makes me feel a type…there are songs that women will be singing, clapping and even dancing and they may be expecting you to be clapping and singing as well [laughs]. When you are not clapping because you don’t identify, it becomes a call for concern. When you sing along for solidarity, it is still a call for concern because it may appear as though you are pregnant as well, of which you are not!” Male #15  “Some men are shy [chuckle] because coming to ANC is like coming into a large pool of women. Perhaps, if is made known that there other men also coming along with their wives, they might assume that they are coming to meet their fellow men” Female #33  The first day I went, I was in a pool of women and all eyes were on me. I was like…OK “what am I doing here?” I am not pregnant! Male #18  “It could also be as a result of bad company or the lack of male models. If a man has bad friends that have not been accompanying their wives for ANC, even if he may want to, the influence of those not doing it will discourage him. He might think if he does it, they may call him names, like he’s being controlled by the wife. He may succumb to peer pressure, not that he likes it. It is just that the desire to identify with and belong in his circle of friends is a deterrent” Female FGD #3  “We still see a pregnant woman like the normal girl who has nothing and can carry on with her duties. But when a baby is born, it is an extra load and we see it as an opportunity to help. It is easier to sacrifice time and come to the hospital when a baby is born because it is challenging for a woman to hold the baby alone immediately after delivery. If she is here and needs to pee, who will she give the baby to? But when she is pregnant, she can come here and do things by herself”  Male FGD #6  “I support what my brother just said. More men come for IWC and not ANC because at that stage, the baby has been dissociated from the mother and it is easier for a man to identify with the baby than during pregnancy.”  Male FGD #1 |
| 4. **Gender, Social & Cultural Norms on Male attendance**  - Male partner sense of superiority about attending ANC - Engendered perception of the time value of ANC attendance - “Woman Wrapper” tag—male fear of losing control and female fear of appearing in control | Men are superior to women and male ANC attendance reduces men to a woman’s level  Not following women for ANC is a male form of maintaining control (Going for ANC with a woman puts men and women on an equal platform)  Men and women cannot sit and receive health education together as though they are equals  Men should be engaged in income-generating activities; ANC attendance is not a good use of their time.  ANC attendance for women is obligatory for women but not for men  Time and the ‘busy’ concept is not the same for men and women  There is a stigma attached to male ANC attendance and men fear the woman wrapper tag which speaks to being emasculated if a man attends ANC. | Capability  Motivation  Opportunity | Self-esteem  Resources  Social norms and comparisons | “I think there is a cultural [gender] attachment to this. You know, we African men we have a certain way of relating with women—it is a kind of boss-subordinate relationship. I am the head of the family. I have to dictate and the woman follows. Men don’t see themselves and women as being equals. So they don’t feel comfortable sitting and being given health education together with women. A man may feel like, ‘if I go to clinic with my wife, it may appear like my wife and I are equal’ ” Male #38  “Yes, pride prevents men from coming because someone will ask that “what is a man doing where there are women, pregnant women, singing and clapping hands?” It is not a man’s thing.” Female #10  “Just to chip in one thing that hinders men from coming….I can say pride and male ego is a barrier to male ANC attendance. Some men think that if you come along for ANC with your wife, she is controlling you” Male FGD #1  “The point is that coming to the clinic for me is a must. Time or no time I must be at the clinic on the appointed date until delivery. That is not the same for my husband. As a man, he must go out and struggle to work so that he can get something (money) to give me for ANC attendance.” Female #19  “For men especially African men, one cannot really condition their activities. Being busy for them would also include the time that they spend in the bars and the time that they hang out with friends It is also in their schedule for the day when they envisage their plans for the day. So, we cannot really try to control them. They are not really flexible enough” Female #11  “Men think that time is money as we usually say so coming here to sit and waste the whole day instead of hustling is not worth it” Male FGD #2  “Another barrier is that the women, …the women themselves…some women say, ‘our husbands have to look for the money, so they cannot come and be here’” Staff FGD #4  “You have the issue of time…time factor which is also something so important because the man is always busy from morning till night… and remember that it is the man who in most cases (I can say about 80% or 70% of cases), the man is the one who provides everything in the house. So, he makes sure that he catches up with those activities in order to be able to provide. Because when he sits somewhere and loses a day, it is really something big that he has lost Male #15  “Some men are shy to accompany their wives to the hospital. Not all men have the courage to feel comfortable in walking behind a woman to go to the clinic” Female #28  “You know others feel that when you come with your wife for ANC, it appears like your wife is controlling you” Female #35  “ You know, African men have a mentality that if they follow their wives for antenatal, people might see them and think that he lacks something to do or he is a “woman wrapper” [weakling, sissy or inadequate man]  “The day we came for ANC, he was the only man who came for ANC and even though some women will think that I am controlling my husband, that was his personal decision” Female #22  “To add to what my brother has said, let me speak from my community. It appears like, attending ANC with a woman is a way of giving up your authority as a man. When your fellow men see you, they look at you like you are a woman [group laughter]. Yes, you are a woman the man because if you were a man, you would not be following your wife to go do women’s things” Male FGD #3 |
| 5. **Health system-related barriers**  - Token-based engagement (ANC claps for men who attend) - Lack of male engagement and female-focused health education - Provider attitude (still testing this theme across data sets) | There is no point in male ANC attendance if clinic staff will focus more on women without engaging men  Token-based engagement by offering men ‘ANC claps’ for showing up in ANC clinics is childish | Opportunity | Organisational culture and climate | “Some men don’t want to be identified among women with claps for ANC attendance. It reduces them to kids.” Female #22  “Some men have said: “I came to the clinic and at the end they said ‘let’s clap for papa, papa came for clinic today’ and they felt like they were in primary school. So they will not come again, because they don’t like being treated as kids”  Staff FGD #3  I blame the ignorance on the part of the health authorities. They have not made a provision for us. If a pregnant woman does not go for antenatal care, even an uneducated grandmother will ask her why because they know it is mandated. However, when a man does not go, nobody will ask him questions because it is not mandated anywhere. I believe that these health institutions should be the ones to make it mandatory and permit men to have their own clinic day or come along with their wives.  Male FGD #7  We know that the ANC register has space for male partners with two phone numbers—one for the person in front of you [pregnant lady] and the contact of her partner. Majority of us providers don’t use the phone numbers for follow up—to ensure that this woman has actually given out the invitation letter or spoken with the partner in question about their visit to the clinic. I think this is a barrier on our part.  Staff FGD #4  “You are the first to bring up this procedure on couple ANC attendance and testing for HIV. That is what we should have done before but the attention of the hospital has mostly been on her.”  Male #31  “During her first ANC visit, I asked her if they said I should come, she said no. That is why I stayed behind”  Male #44 |
| 6. **Emotional reactions to male ANC attendance**  - Fear of being judged or discovered for extra-marital affairs - Avoidance of responsibility - Fear of getting tested for HIV | Pregnancy is perceived as a time for extra-marital experimentation and some men don’t want to be discovered through ANC attendance.  The fear of having health education echo on the minds and consciences of most men who are unfaithful to their partners prevent them from attending ANC.  Attending ANC is an explicit form of taking financial responsibility which some men want to shy away from  Antenatal attendance comes with the requirement for couple testing and some men are not ready for testing.  Associating ANC attendance with couple HIV testing is a deterrent.  The prospect of attending ANC floods both men and women with a range of negative emotions like fear and shyness | Motivation | Fear  Negative emotions | “Once a woman becomes pregnant, some  men get into the practice of what we call ‘sidechicks’ ” Female #2  “Some men avoid ANC all-together because they don't want to be judged about their sinful lives and extra-marital affairs. They don't want health talks at the clinic to echo in their mind and their consciences” Female #11  “Some men are ashamed especially when they have impregnated many girls in the neighbourhood and they don’t want to be tagged as a particular woman’s husband when there are other women he has impregnated and he is denying being responsibility for their pregnancies.” Female #32  “The ANC testing requirement is also a barrier. Perhaps they are aware that if they come, they will be checked and tested and for those who are unaware/unsure of their status, they don’t want to come”  Male #45  “If a woman is coming for ANC and she tells her husband that they are going to do HIV testing for both of them, at that point you will hear the man say ‘my coming is not necessary’. The name HIV alone cancels the whole issue” Female #20  “I think there is also a financial barrier. Some men don’t want to attend ANC because attending will mean they are taking responsibility as authors of the pregnancy and this also means they are required to show up as fathers and engage in financial responsibilities”  Female #38  “Contextually we live in a society that already places that demand on the man to take responsibility of the pregnancy and to follow up materially and emotionally…”  Staff FGD #1 |
| 7. **Women are barriers**  - Maternal extortion - Maaaternal discomfort | Women use pregnancy as an opportunity to extort money from their partners and see male ANC attendance as a barrier to their financial scheme.  Some women are barriers as ANC is a means for them to raise extra finances  Women are shy on behalf of their partners who are required to sit with other women for intimate discussions on birth and pregnancy |  |  | “Women are also barriers. They don’t want their partners to come as participant number 5 mentioned because they don’t want the men to know the amount they are spending for ANC. They use pregnancy as a forum to exhort a lot of money from their partners.”  Staff FGD #8  Some women don’t also give the opportunity for their husbands to come with them for financial reasons. They don’t want them to know what is happening here and how much is being spent. Female #40 |
