## Supplementary material for "“Go and bring your husband”: a COM-B guided qualitative study on the barriers to male involvement in antenatal care in Bamenda Health District, Cameroon": Conceptual analysis

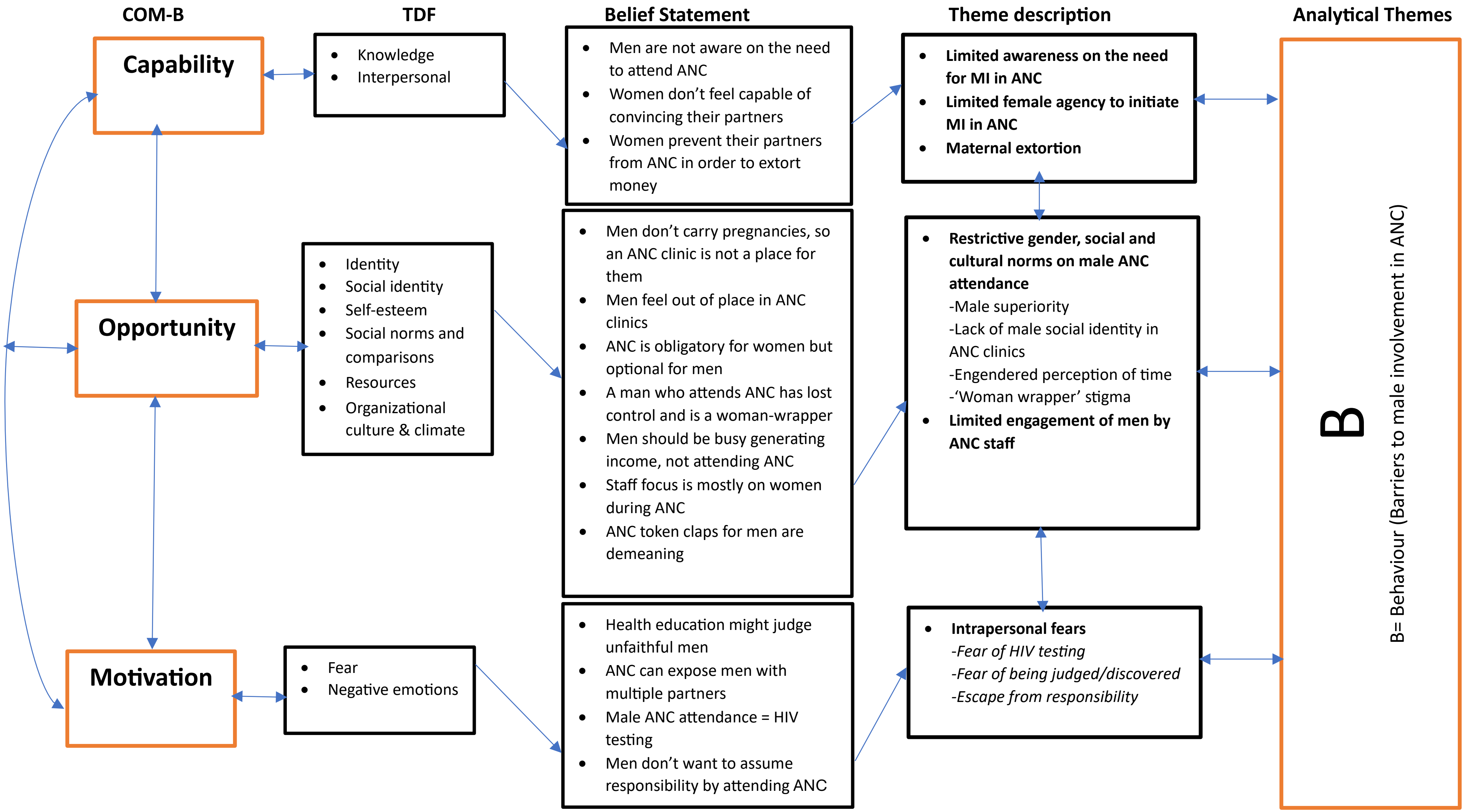


Figure 1: Conceptual analysis of barriers to male involvement underpinned by the COM-B model of behaviour change (29,31)
